# Genome-Wide Meta-Analysis Identifies Genetic Risk Loci for Mono- and Polyneuropathies in 983,477 Individuals

**DOI:** 10.1101/2025.08.06.25333006

**Authors:** Martin Broberg, FinnGen, Eija Kalso, Hanna M. Ollila

**Author notes:** See detailed contributor list in Table S1 (FinnGen).

## Abstract

Peripheral neuropathies are common neurological disorders affecting sensory, autonomic, and motor nerves, with an estimated prevalence exceeding 2% in the general population. Typical symptoms include numbness and distal limb muscle weakness, resulting from somatosensory nerve damage. Here, we investigate the genetic architecture of mono- and polyneuropathies and their relationships with comorbid traits using data from FinnGen and the UK Biobank. Our genome-wide association study (GWAS) and meta-analysis identified 48 genome-wide significant (p < 5 × 10^−8^) independent loci and 66 fine-mapped signals. These included associations with genes involved in neurotransmitter signaling (*HTR3A*), immune function (*HLA-DQB1*, *BCL11A*), extracellular matrix remodeling (*COL11A1*, *ADAMTS17*, *LOXL4*), axon guidance and neural development (*DCC*, *ETV1*, *NEGR1*), and carpal tunnel syndrome (*DIRC3*). Phenome-wide association studies (PheWAS), genetic correlation, and Mendelian randomization analyses supported shared genetic links with sleep problems, chronic pain, and psychiatric disorders. Together, our results highlight a strong polygenic basis for neuropathies and confirm their complex comorbid relationships with sleep, pain, psychiatric, and autoimmune traits.

**Author approval:** All authors have seen and approved the manuscript.

## Introduction

Neuropathies are common and affect over 2% of individuals within the general population (1) and even a larger proportion, approximately 7%, in the elderly (2,3). Furthermore, neuropathic pain is caused by damage of the somatosensory nerves, affecting 3-8% of the general population (1,2,4). Typically, neuropathies associate with an underlying disease such as diabetes, hypothyroidism, immunological diseases, traumas or toxic agents. In addition, chemotherapy is often observed as a common causative factor. However, in several cases, neuropathies are considered idiopathic as the pathophysiology, or the causative factors cannot be identified or are not known. Neuropathic symptoms include sensory and autonomic symptoms and muscle weakness, which most frequently manifest in the distal limbs (3,5) and facial area. Sensory symptoms include increased or decreased sensitivity to various noxious and innocuous stimuli that can be of thermal, mechanical, and chemical stimulus.

The first line pharmacological treatment of neuropathic pain includes low dose-tricyclic and SNRI-type antidepressants, and gabapentinoids (pregabalin and gabapentin). Neuropathic pain does not respond to non-steroidal anti-inflammatory drugs as nociceptive pain does (4,6).

Neuropathies do not inevitably follow nerve injury, as evidenced by their absence in some diabetic and chemotherapy-treated patients (4). Comorbidities associated with neuropathies include autoimmune conditions, (7), asthma (8), sleep problems (9,10), depression, and mood disorders (4,11,12). Moreover, the relationship between comorbidities can be causal. For example, recent findings from our group established genetic and causal links between acute or chronic pain and insomnia (13).

Neuropathies and their comorbid pain have also been reported to be associated with sleep disorders, depression and anxiety (4,12,14). There are multiple genes that have been reported as associated with neuropathic pain, including *IL10* encoding an anti-inflammatory cytokinin (15), *PRPH* encoding a cytoskeletal protein in the peripheral nervous system (16), *CEP72* encoding a leucine-rich repeat protein on the centrosome (17), *VAC14* encoding a scaffold protein part of the PIKfyve complex (18), *IL2RA* encoding the interleukin receptor-2 alpha subunit (19) and *XIRP2* which is involved with actin cytoskeleton organization (20).

Earlier work, including genome-wide association studies, have demonstrated a robust genetic component with pain and neuropathies (1,3,6). Earlier GWASs have identified a variant near mitochondrial copper sulfate transporter *SLC25A3* gene (rs369920026) as a risk factor for neuropathic pain (21). Moreover, GWAS of polyneuropathies showed an association at *SNX8* locus (rs10950641), *PCP2*, (rs6796803), *KNG1* (rs6796803) and *RORA* (rs4775319) (22), and with *B4GALNT3* (rs7294354) and *NR5A2 (*rs147738081(23). Together these earlier findings show a consistent polygenic signal with neuropathies.

Understanding the biological mechanisms that lead to neuropathic pain or protect a subset of individuals from neuronal damage or severe symptoms would benefit a substantial proportion of the population or provide novel pharmaceutical targets. To understand the pathophysiological mechanisms of neuropathies for improved diagnosis and treatment, there is a need to explore this component in more detail.

Here, explored the biological mechanisms that lead to neuropathies. We focused on mono and polyneuropathies and used a genome-wide association study (GWAS) approach using FinnGen and the UK Biobank (UKB) research projects.

In addition to GWASs, we compared the data to other GWASs of neuropathies and studied the causal relationship of neuropathies to sleep problems and psychiatric factors by utilizing the post-GWAS methods PheWas, eQTL analysis, HLA fine-mapping, colocalization, genetic correlation and Mendelian randomization (MR). We discover 48 genetic signals for neuropathies and a strong polygenic signal that both clarifies underlying disease mechanisms and may facilitate development of pharmaceuticals for neuropathies and neuropathic pain.

## Materials and Methods

### FinnGen

FinnGen is a large-scale research project where participants are recruited from hospitals as well as prospective and retrospective epidemiological and disease-based cohorts. The participants are of Finnish ancestry, and all have been genotyped with genome-wide arrays. The genotype data are combined with longitudinal registries that record phenotypes and health events (including ICD-based diagnosis) over the entire lifespan including the Care Register for Health Care (inpatient and outpatient), Causes of Death Registry, the National Infectious Diseases Registry, Cancer Registry, Primary Health Care Registry (outpatient) and Medication Reimbursement Registry. This study used data from FinnGen R12 (N = 500,348 individuals). For this study, we used the registered ICD-10 diagnoses as recorded across the cohort.

### Genotyping and imputation in FinnGen

FinnGen genotype data is based on an imputation reference panel of 3,775 Finnish individuals that have been whole genome-sequenced at a 25–30x resolution (24). The panel contains a total of 16,962,023 single nucleotide polymorphisms, and insertions and deletions (minor allele count of ≥3). After QC based on an imputation information (INFO) score of >0.6, 16,387,711 of the variants remain for further genetic analyses. As of FinnGen Release 12, a total of 500,348 have been genotyped using a custom Axiom FinnGen1 array (24).

### Neuropathic pain phenotype in FinnGen

The following ICD-10 codes were used to define poly and mono neuropathies, where any lifetime diagnosis for an individual was then deemed a case; G56, G57, G58.0, G58.7, G58.8, G589, G59, G60, G61.1, G61.8, G61.9, G620, G62.1, G62.2, G62.8, G62.9 and G63.

### GWAS in FinnGen

The GWAS for neuropathies was performed using REGENIE on the FinnGen R12 cohort (cases = 58,967, controls = 441,381, Table S2) using Finnish participants, where over 70% had at least one 3^rd^ degree relative or closer, within the cohort (24,25). The covariates used for the analysis included: sex, imputed age, age at death or end of follow up, 10 principal components and microarray genotyping batch. The default REGENIE filtering of a minimum allele count > 5 was used, along with the firth regression option. As HLA alleles are heavily associated with diabetes, BMI, and neuropathies, as a sensitivity analysis, we repeated the GWAS in FinnGen but removed individuals with any diagnosis of diabetes-linked neuropathy (ICD-10 codes E10.4 and E11.4) from the analysis. Thus, the diabetic sensitivity GWAS included 57,340 cases and 439,658 controls.

### FinnGen ethics statement

Study subjects in FinnGen provided informed consent for biobank research, based on the Finnish Biobank Act. Alternatively, separate research cohorts, collected prior the Finnish Biobank Act came into effect (in September 2013) and start of FinnGen (August 2017), were collected based on study-specific consents and later transferred to the Finnish biobanks after approval by Fimea (Finnish Medicines Agency), the National Supervisory Authority for Welfare and Health. Recruitment protocols followed the biobank protocols approved by Fimea. The Coordinating Ethics Committee of the Hospital District of Helsinki and Uusimaa (HUS) statement number for the FinnGen study is Nr HUS/990/2017.

The FinnGen study is approved by Finnish Institute for Health and Welfare (permit numbers: THL/2031/6.02.00/2017, THL/1101/5.05.00/2017, THL/341/6.02.00/2018, THL/2222/6.02.00/2018, THL/283/6.02.00/2019, THL/1721/5.05.00/2019 and THL/1524/5.05.00/2020), Digital and population data service agency (permit numbers: VRK43431/2017-3, VRK/6909/2018-3, VRK/4415/2019-3), the Social Insurance Institution (permit numbers: KELA 58/522/2017, KELA 131/522/2018, KELA 70/522/2019, KELA 98/522/2019, KELA 134/522/2019, KELA 138/522/2019, KELA 2/522/2020, KELA 16/522/2020), Findata permit numbers THL/2364/14.02/2020, THL/4055/14.06.00/2020, THL/3433/14.06.00/2020, THL/4432/14.06/2020, THL/5189/14.06/2020, THL/5894/14.06.00/2020, THL/6619/14.06.00/2020, THL/209/14.06.00/2021, THL/688/14.06.00/2021, THL/1284/14.06.00/2021, THL/1965/14.06.00/2021, THL/5546/14.02.00/2020, THL/2658/14.06.00/2021, THL/4235/14.06.00/2021, Statistics Finland (permit numbers: TK-53-1041-17 and TK/143/07.03.00/2020 (earlier TK-53-90-20) TK/1735/07.03.00/2021, TK/3112/07.03.00/2021) and Finnish Registry for Kidney Diseases permission/extract from the meeting minutes on 4th July 2019.

The Biobank Access Decisions for FinnGen samples and data utilized in FinnGen Data Freeze 12 include: THL Biobank BB2017_55, BB2017_111, BB2018_19, BB_2018_34, BB_2018_67, BB2018_71, BB2019_7, BB2019_8, BB2019_26, BB2020_1, BB2021_65, Finnish Red Cross Blood Service Biobank 7.12.2017, Helsinki Biobank HUS/359/2017, HUS/248/2020, HUS/430/2021 §28, §29, HUS/150/2022 §12, §13, §14, §15, §16, §17, §18, §23, §58, §59, HUS/128/2023 §18, Auria Biobank AB17-5154 and amendment #1 (August 17 2020) and amendments BB_2021-0140, BB_2021-0156 (August 26 2021, Feb 2 2022), BB_2021-0169, BB_2021-0179, BB_2021-0161, AB20-5926 and amendment #1 (April 23 2020) and it’s modifications (Sep 22 2021), BB_2022-0262, BB_2022-0256, Biobank Borealis of Northern Finland_2017_1013, 2021_5010, 2021_5010 Amendment, 2021_5018, 2021_5018 Amendment, 2021_5015, 2021_5015 Amendment, 2021_5015 Amendment_2, 2021_5023, 2021_5023 Amendment, 2021_5023 Amendment_2, 2021_5017, 2021_5017 Amendment, 2022_6001, 2022_6001 Amendment, 2022_6006 Amendment, 2022_6006 Amendment, 2022_6006 Amendment_2, BB22-0067, 2022_0262, 2022_0262 Amendment, Biobank of Eastern Finland 1186/2018 and amendment 22§/2020, 53§/2021, 13§/2022, 14§/2022, 15§/2022, 27§/2022, 28§/2022, 29§/2022, 33§/2022, 35§/2022, 36§/2022, 37§/2022, 39§/2022, 7§/2023, 32§/2023, 33§/2023, 34§/2023, 35§/2023, 36§/2023, 37§/2023, 38§/2023, 39§/2023, 40§/2023, 41§/2023, Finnish Clinical Biobank Tampere MH0004 and amendments (21.02.2020 & 06.10.2020), BB2021-0140 8§/2021, 9§/2021, §9/2022, §10/2022, §12/2022, 13§/2022, §20/2022, §21/2022, §22/2022, §23/2022, 28§/2022, 29§/2022, 30§/2022, 31§/2022, 32§/2022, 38§/2022, 40§/2022, 42§/2022, 1§/2023, Central Finland Biobank 1-2017, BB_2021-0161, BB_2021-0169, BB_2021-0179, BB_2021-0170, BB_2022-0256, BB_2022-0262, BB22-0067, Decision allowing to continue data processing until 31st Aug 2024 for projects: BB_2021-0179, BB22-0067,BB_2022-0262, BB_2021-0170, BB_2021-0164, BB_2021-0161, and BB_2021-0169, and Terveystalo Biobank STB 2018001 and amendment 25th Aug 2020, Finnish Hematological Registry and Clinical Biobank decision 18th June 2021, Arctic biobank P0844: ARC_2021_1001.

### The UK Biobank

The UK Biobank (UKB) consists of over 500,000 participants aged between 40-69 years, who were assessed between 2006-2010 across 22 assessment centers in the UK. The assessment was based on informed consent, and consisted of an interview phase, filling out a touch screen questionnaire, blood, saliva, and urine sampling for analysis, and physical and functional measurements. Longitudinal diagnostic data were provided from health care records (Hospital in-patient and Primary care). This study is based on UKB Application 22627.

### Genotyping and imputation in UKB

The UK biobank genotype data is based on a custom Axiom array by Affymetrix and the UK BiLEVE Axiom array, where both arrays have a 95% marker overlap (26). A total of 812,428 markers were used in the array to genotype 489,212 individuals. Imputation was then performed using a combination of the 1000 Genomes phase 3, UK10K and the Haplotype Reference Consortium datasets as reference panels. An INFO score of >0.3 was used for additional QC and a minor allele frequency (MAF) of > 0.001 (26).

### Neuropathic pain phenotype in UKB

The following ICD-10 codes were used to define poly and mono neuropathies, where any lifetime diagnosis for an individual was then deemed a case; G56, G57, G58.0, G58.7, G58.8, G589, G59, G60, G61.1, G61.8, G61.9, G620, G62.1, G62.2, G62.8, G62.9 and G63.

### GWAS in UKB

We performed a secondary GWAS of general CHD using UKB (cases = 20,600, controls = 466,528, Table S2) using the REGENIE software (25), minimum allele count 3 and using sex, age, PC1-10, array used for SNP detection, and assessment center as covariates.

### Meta-analysis

We performed a GWAS meta-analysis using METAL (sample size based analysis) (27), combining the two GWAS datasets for a combined N = 983,477 (Table 3) and further extracting only SNPs that were detected by REGENIE in both GWASs. The lead loci in the independent GWASs and the meta-analysis were identified using a combined approach of the topr R package v2.0.0 (28) and LocusZoom v0.14 (29). We also tested geneset enrichment in the FUMA platform for the lead loci identified in the meta-analysis (30).

### PheWas analyses across cohorts

We utilized the available PheWas datasets provided by the UKB, FinnGen, Biobank Japan (31) and VA Million Veteran Program (32), specifically looking at the associations between the genomic loci in *DIRC3* and *HLA-DQB1* and non-diabetic neuropathic pain, neuronal and connective tissue disorders, and related neurological comorbidities.

### HLA fine-mapping in FinnGen

We fine-mapped genome-wide significant signals in the MHC area of chromosome 6 using logistic regression modelling on the imputed HLA alleles in FinnGen R12. The FinnGen HLA imputation is done using the HIBAG R library and a customized training panel of Finnish individuals (33,34). Logistic regression was performed using the glm function in R v4.4.0 with age, sex and principal components 1-10 as covariates.

### eQTL and pQTL association search

To find whether the genome-wide significant SNPs from the meta-analysis GWAS could have an effect on the expression of nearby genes, we utilized the FinnGen annotation tool (https://anno.finngen.fi) which provides gene/variant associations from the eQTL catalogue (35) and the GTEx portal database (www.gtexportal.org) (36,37), the UKB pQTL 3K dataset (38), deCODE pQTL 2021 dataset (39) and in-house FinnGen datasets.

### Fine-mapping and colocalization

For fine-mapping the meta-analysis results we used the SuSiE (40) v0.14.2 R package, with the FinnGen samples for creating the LD-matrix for the analysis. The resulting fine-mapped regions were subsequently used with the coloc v5.2.3 (41,42) R package, against the GTEx V8 eQTL dataset (43), EMBL-EBI eQTL catalogue dataset (44), FUSION, UK Biobank Finucane biomarkers (45), Kolberg immune cell microarray data (46), and FinnGen in-house datasets.

### Genetic correlation

To understand the overall genetic overlap between neuropathies and traits relating to pain, sleep and psychiatric conditions, genetic correlation was performed using the LDSC software (47) v1.0.1 and the 1000G European phase 3 LD reference panel. We compared the neuropathies trait to other cohorts as well as against traits within FinnGen R12.

### Mendelian randomization

Based on the genetic correlation, we chose 15 general, psychiatric, sleep, and substance abuse traits for MR analysis to further understand their causality. These traits included: Parkinson’s disease, Alzheimer’s disease, attention-deficit/hyperactivity disorder (ADHD), post-traumatic stress disorder (PTSD), alcohol use disorder (AUD), weekly alcohol consumption, bipolar disorder, body mass index (BMI), diabetes, parental longevity, insomnia, anxiety, depression, neuroticism, schizophrenia, and multisite chronic pain). The MR analysis was performed using the TwoSampleMR (48) R-package. There are multiple methods for MR analysis. In this study, we focused on the inverse-variance weighting (IVW) method. The IVW method produces a weighted regression-averaged ratio estimate of the exposure instruments to the outcome to calculate an overall causal estimate.

## Results

### Meta-analysis identifies 48 genetic loci

To understand the genetic architecture of neuropathic pain, we performed a meta-analysis from the UKB and FinnGen neuropathic pain GWAS, including a total of 79,567 cases and 907,909 controls (**Table S2**). The meta-analysis combining both datasets identified 48 genome-wide significant loci (**Table S3**, Figure 1). Notably, the lead variants are located at the *HTR3A* region and encode the 5-HT3A serotonin receptor and potentially highlight serotonergic mechanism through 5-HT3A in pain modulation. Other genes included also extracellular matrix remodeling (*COL11A1*, *ADAMTS17*, *LOXL4*), axon guidance and neural development (*DCC*, *ETV1*, *NEGR1*), and immune regulation (*HLA-DQB1*, *BCL11A*). Mechanosensation and ion channel activity are represented by *PIEZO1* and *KCNT2*, suggesting roles in neuronal excitability.

**Figure 1.**
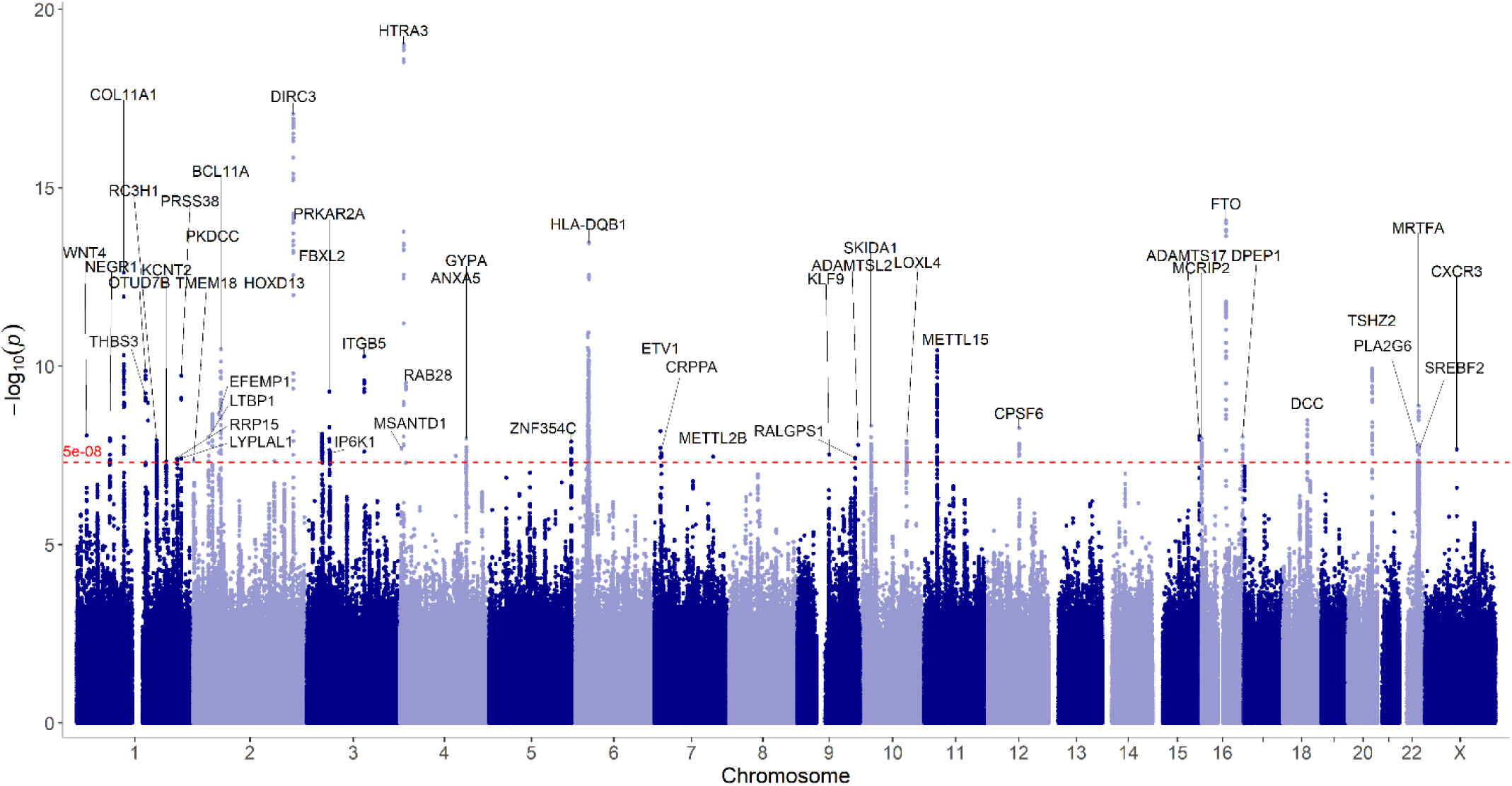
Meta-analysis of neuropathies in FinnGen and UKB detects 48 lead variants. Manhattan plot of the meta-analysis results made based on the topr R package, indicating the nearest curated gene to the lead variant locations.

**Figure 2.**
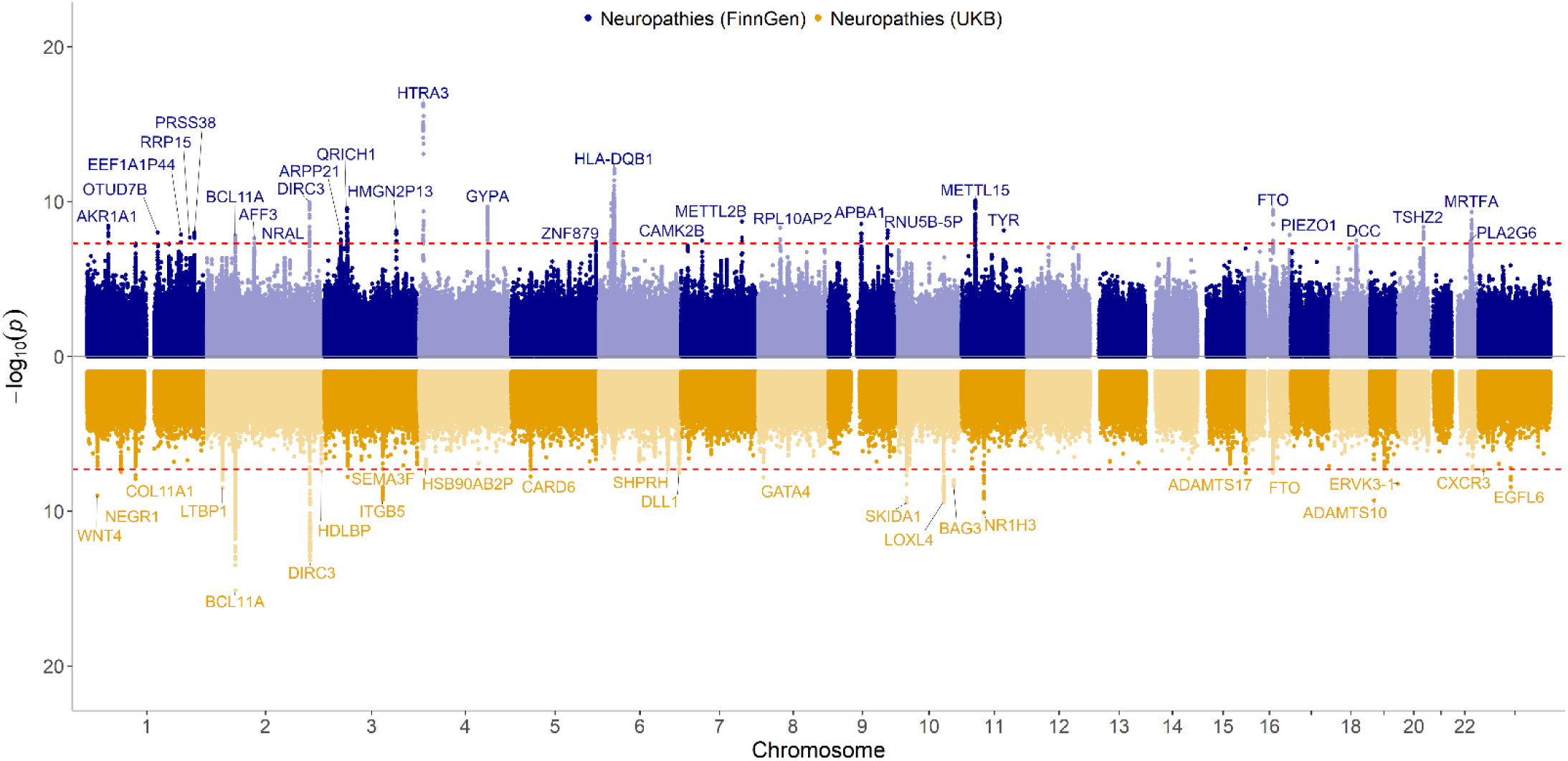
GWASs of mono and polyneuropathies in FinnGen and UKB elucidate multiple genome-wide significant loci. Miami plot of FinnGen (blue color, upper) and UKB (gold color, lower) GWAS summary statistics, genome-wide significant loci are named by the nearest curated gene. The dashed red line indicates genome-wide significant -log_10_(p) = 7.3.

The associating variants were robust across data sets with majority the loci showing consistent effect size association both in UKB and in FinnGen (**Table S4-5**). Moreover, 29 genetic loci in FinnGen and 24 genetic loci in UKB were significant in the respective cohorts (Table S4 and 5, Figure 1). Two of the loci; *DIRC3* and *FTO*, were genome-wide significant in both cohorts.

### Neuropathic pain loci are shared with other traits

To further elucidate contributing factors in neuropathic pain we examined if individual genetic loci are enriched for other diagnoses. To do this we performed gene set enrichment analysis with FUMA using the 48 genomic loci. We identified 91 significantly (adjusted p-value < 0.05) enriched gene sets with earlier reported GWAS catalog genes (**Table S6**). These included carpal tunnel syndrome, BMI, obesity, neuroticism, bone density and cognitive ability.

### Fine-mapping, colocalization and eQTL analyses

Following the meta-analysis, we examined if the lead variants observed in the meta-analysis also associated with expression changes of nearby genes. We observed strong expression QTL signal with majority of the lead genetic loci with 13,193 overall eQTLs or pQTLs across tissues (**Table S7**). Additionally, the results of the SuSiE fine-mapping produced 66 credible (posterior probability > 0.95) sets (**Table S8**) and colocalization using the fine-mapping regions (**Table S9**), demonstrated 122 significant colocalizations (posterior probability > 0.95). From the combined eQTL and pQTL analysis, *DIRC3* locus demonstrated the most significant association (eQTL catalogue dataset ID: QTD000076, *DIRC3-AS1* beta = 0.44, p-value = 5.94×10-19, Table S7.

### Nine HLA alleles associated with neuropathies

The results from the HLA allele finemapping in FinnGen demonstrated nine HLA alleles significantly associated with neuropathies, of which the top five all are part of a well-established 8.1 ancestral haplotype (49) (**Table 1**).

**Table 1.** Nine HLA alleles with significant Bonferroni corrected p-values (p < 0.05) against neuropathies.

| **Allele** | **beta** | **se** | **p** | **Bonferroni p** |
| --- | --- | --- | --- | --- |
| B*08:01 | 0.07 | 0.01 | 1.15E-07 | 2.14E-05 |
| DQB1*02:01 | 0.06 | 0.01 | 4.61E-06 | 8.57E-04 |
| DRB1*03:01 | 0.06 | 0.01 | 5.20E-06 | 9.67E-04 |
| DQA1*05:01 | 0.06 | 0.01 | 7.16E-06 | 1.33E-03 |
| C*07:01 | 0.05 | 0.01 | 8.31E-06 | 1.55E-03 |
| DQA1*03:01 | 0.05 | 0.01 | 2.12E-05 | 3.94E-03 |
| DQB1*03:02 | 0.05 | 0.01 | 2.28E-05 | 4.24E-03 |
| DPB1*01:01 | 0.07 | 0.02 | 8.73E-05 | 1.62E-02 |
| DPB1*04:02 | -0.04 | 0.01 | 1.84E-04 | 3.42E-02 |

### PheWas searches demonstrate significant associations between rs11676136 and connective tissue diseases

As sleep, psychiatric traits, chronic pain, autoimmunity and pulmonary infections are commonly associated with neuropathies, we assessed the phenome-wide association of all lead variants in other traits. We observed association of the 48 lead variants at genome-wide significant level with 6,703 diseases and phenotypes. Notably, autoimmune traits and sleep traits were also observed in this analysis (**Table S10**). Additionally, we tested the finemapping/colocalization *DIRC3* locus lead SNPs rs11676136 specifically in the Million Veteran Program, Japan Biobank and UKB datasets selecting genome-wide (<5×10^-8^) associations. For rs11676136 we found significant associations with mixed connective tissue disease (p value = 6.85×10^-11^, beta = -0.01) and basal ganglia structure (p value = 1.00×10^-8^, beta = N/A) in the UKB and synovitis in the VA million veteran program meta-analysis (p value = 1.1×10^-14^, beta = -0.1).

However, in the FinnGen PheWas, disregarding endpoints with overlapping ICD-10 codes, we found significant associations for *DIRC3* with autoimmune diseases (beta = -0.03, p-value = 5.5×10^-10^), trigger finger (beta = -0.22, p-value = 2.0×10^-10^) and autoimmune hypothyroidism (beta = -0.083, p-value = 1.6×10^-30^).

### Neuropsychiatric, sleep and pain traits are connected with neuropathic pain

To study the genetic associations between neuropathies and comorbid traits, we tested the genetic correlation between neuropathies and these traits (**Table 2**) and further studied their causal linkage using MR (**Table 3**). Furthermore, we also performed genetic correlation within FinnGen using 2,469 FinnGen R12 GWAS datasets. This analysis demonstrated significant (Bonferroni adjusted p-value < 0.05) correlation between neuropathies and 462 traits, including carpal tunnel syndrome (p-value < 10^-244^), and nerve/nerve root/plexus disorders (p-value < 10^-244^), sleep apnoea (p-value = 5.1×10^-38^), antidepressant use (1.4×10^-37^, **Table S11**).

**Table 2.** Neuropathies show significant correlation with sleep, psychiatric, and pain traits.

| Neuropathies vs | RG | SE | P-value | Bonferroni p-value |
| --- | --- | --- | --- | --- |
| Insomnia | 0.28 | 0.027 | $1.7 \times 10^{-25}$ | $3.0 \times 10^{-24}$ |
| Short sleep | 0.34 | 0.033 | $1.7 \times 10^{-24}$ | $3.0 \times 10^{-23}$ |
| Anxiety | 0.29 | 0.041 | $1.2 \times 10^{-12}$ | $2.2 \times 10^{-11}$ |
| Depression | 0.31 | 0.026 | $7.6 \times 10^{-34}$ | $1.4 \times 10^{-32}$ |
| Sleep duration (Accelerometer) | 0.14 | 0.065 | $3.2 \times 10^{-2}$ | $5.8 \times 10^{-1}$ |
| Mood instability | 0.29 | 0.033 | $1.2 \times 10^{-18}$ | $2.2 \times 10^{-17}$ |
| Schizophrenia | -0.15 | 0.025 | $6.1 \times 10^{-9}$ | $1.1 \times 10^{-7}$ |
| Long sleep | 0.1 | 0.039 | $8.1 \times 10^{-3}$ | $1.5 \times 10^{-1}$ |
| Short sleep | 0.33 | 0.033 | $6.8 \times 10^{-24}$ | $1.2 \times 10^{-22}$ |
| Chronotype continuous | 0.094 | 0.022 | $2.6 \times 10^{-5}$ | $4.7 \times 10^{-4}$ |
| Chronotype binary | 0.063 | 0.021 | $2.0 \times 10^{-3}$ | $3.6 \times 10^{-2}$ |
| Neuroticism | 0.18 | 0.025 | $1.7 \times 10^{-12}$ | $3.1 \times 10^{-11}$ |
| Restless legs syndrome | 0.2 | 0.067 | $3.2 \times 10^{-3}$ | $5.8 \times 10^{-2}$ |
| Severe covid19 | 0.17 | 0.062 | $5.6 \times 10^{-3}$ | $1.0 \times 10^{-1}$ |
| Fibromyalgia | 0.46 | 0.039 | $1.1 \times 10^{-31}$ | $2.0 \times 10^{-30}$ |

**Table 3.** Two-sample MR results using FinnGen R12 neuropathies instruments as exposure or as outcome.

| **Outcome: Neuropathies** |  |  |  |
| --- | --- | --- | --- |
| **Exposure** | **nSNPs** | **OR [95% CI]** | **P (IVW)** |
| Insomnia | 224 | 1.13[1.10-1.17] | $1.4 \times 10^{-14}$ |
| Chronotype | 276 | 0.91[0.85-0.97] | $6.3 \times 10^{-3}$ |
| Anxiety | 99 | 1.05[1.01-1.09] | $5.9 \times 10^{-3}$ |
| MCP | 61 | 2.27[1.94-2.66] | $3.7 \times 10^{-24}$ |
| **Exposure: Neuropathies** |  |  |  |
| **Outcome** | **nSNPs** | **OR [95% CI]** | **P (IVW)** |
| Neuroticism | 18 | 1.04[0.93-1.18] | 0.038 |

The two-sample MR analysis using FinnGen R12 as the Neuropathies instrumental variables revealed, similarly to previous pain-sleep/psychiatric traits, risk factors including insomnia, anxiety, multi-site chronic pain (MCP). Differently from the general pain phenotype which included neuropathic pain categories neuralgias and phantom pain, but not neuropathies.

### Genetic association with neuropathic pain and diabetes

As neuropathies can co-occur or be caused by comorbid diseases like diabetes, we examined the genetic associations in population free from diabetes. The sensitivity analysis removing all individuals with neuropathies stemming from type I or type II diabetes from the analysis in FinnGen (ICD-10 codes E10.4 and E11.4) yielded 38 genome-wide significant associations (**Table S12**) including a genome-wide significant loci in the MHC region at SNP rs2050189 (p-value = 2.31×10^-8^, beta = 0.043, SE = 0.0078) in the *TSBP1* gene, 286,791 bp upstream of the *HLA-DQB1* lead variant. The *DIRC3* loci shifted 3,296 bp downstream to the nearest genome-wide SNP rs2113825 (p-value = 4.1×10^-10^, beta = -0.048, SE = 0.0077). In addition, 14 of the gene-based loci associated with neuropathic pain in the main analysis showed consistent association (**Table S12**).

## Discussion

Here we performed a GWAS-based study of neuropathies in two cohorts, FinnGen and UKB. Our meta-analysis identified 48 genome-wide significant (p < 5×10-8) independent genetic loci implicated in immunity, DNA binding, receptor signaling, and growth factor pathways. Additionally, we found significant eQTLs support for several genomic loci. Furthermore, based on PheWas data across UKB, FinnGen, Biobank Japan, and the VA Million Veteran Program, we found genome-wide significant associations of sleep, psychiatric, pain and autoimmune both at the level of individual genes, at level of single variants and with formal enrichment of genes. Overall, these results highlight the polygenic nature of chronic pain with 48 loci and 66 fine-mapped signals and highlight novel biology at neuropsychiatric and immune traits.

We observed the strongest association at the *HTR3A* region. *HTR3A* encodes for the 5-HT3A serotonin receptor and has been suggested as a target for pain management in earlier studies (50,51). Overall, this finding highlights a possible serotonergic mechanism in neuropathic pain. Other genes cluster around extracellular matrix remodeling (*COL11A1*, *ADAMTS17*, *LOXL4*), axon guidance and neural development (*DCC*, *ETV1*, *NEGR1*), and immune regulation (*HLA- DQB1*, *BCL11A*). Mechanosensation and ion channel activity are represented by *PIEZO1* and *KCNT2*, suggesting roles in neuronal excitability. All these genes, and the broader genomic region deserve careful follow-up in functional models. Such experiments may elucidate novel biology in neuropathic pain.

Comparing our results with previous genetic studies relating to neuropathies, the nearest overlapping loci was with *PCP2* (22), which is approximately 1 Mbp upstream of the genome-wide significant loci rs62621197 found here. However, this study was significantly smaller in size (cases = 130, controls = 913) and focusing on cancer patients. The most similar GWAS dataset, by Winsvold et al. (23), which focused on idiopathic neuropathies using the ICD-10 codes G60.3, G60.9 and G62.9 had 2,093 cases and 445,456 controls. We used a wider category for mono- and polyneuropathies which included the ICD-10 codes G56, G57, G58.0, G58.7, G58.8, G589, G59, G60, G61.1, G61.8, G61.9, G620, G62.1, G62.2, G62.8, G62.9 and G63. This was to capture a more comprehensive view of the genetic architecture behind neuropathies.

The MHC region housing the HLA alleles is a complex region in the human genome with one of the highest levels of gene density, polymorphisms, and linkage disequilibrium (LD) (52–54), and thus the specific eQTLs, effect sizes and PheWas associated with the *HLA-DQB1* loci described here can be difficult to interpret as specifically associated with that gene. Furthermore, this locus is heavily associated with diabetes (55,56), an established comorbidity of neuropathies (20,57). However, the HLA finemapping performed in FinnGen, along with the sensitivity GWAS removing individuals diagnosed with neuropathies from type 1 and type 2 diabetes, still demonstrated a significant association of the *MHC* region with neuropathies, suggesting a robust association between *HLA* and neuropathies. This link is accentuated by the HLA fine-mapping result implicating several members of the 8.1 ancestral haplotype, which is described as associated with autoimmunity (49). The results suggest that for a subset of the signals, the contributing comorbidities such as type I or type II diabetes likely modify the genetic association with the neuropathy, in a limited way. Furthermore, the PheWas data demonstrated associations of *HLA-DQB1* and non-diabetic comorbidities of neuropathies.

Here, the fine-mapping and colocalization combination indicated *DIRC3* locus as significantly colocalized with other datasets. The lead variant at the *DIRC3* locus is an intergenic SNP, closest to the *DIRC3-AS1* or *DIRC3* genes. The exact roles of *DIRC3-AS1* and *DIRC3* are unknown, but *DIRC3* encodes a long non-coding RNA, and is primarily associated with carcinomas and cancer (58,59). The *DIRC3* lead signal highlighted in the fine-mapping, colocalization and eQTL analyses demonstrates accentuates the strong connections between neuropathies and neuronal and connective tissue disorders (60). Additionally, *DIRC3* has previously been demonstrated to be associated with trigger finger in a UKB GWAS, and colocalized with carpal tunnel syndrome (61). The *DIRC3* gene affects the transcription of *IGBP5* and their pathway has been suggested to be a potential pharmacological target in treatment of carpal tunnel disorder and trigger finger (61).

Carpal tunnel syndrome itself is commonly an outcome of neuropathy (62). The connection between carpal tunnel syndrome, trigger finger, hypothyroidism, arthritis and diabetes are underlined in our study, and the results suggests that this gene could be considered for further investigation of other types of neuropathies as well. Interestingly, our eQTL, pQTL, fine-mapping and colocalization suggested that brain tissue was the most significantly associated tissue type with rs11676136 and *DIRC3-AS1*/*DIRC3* expression. This provides grounds for further exploration of the role of *DIRC3* in brain tissue or neurons in general.

The MR and genetic correlation analyses both support the links between neuropathies and sleep problems and psychiatric traits and other traits with known neuropathic associations (asthma, restless legs, infection).

## Strength and limitations

Using a large sample, we were able to gain sufficient statistical power to detect genetic risk loci, leading to the identification of 48 genome-wide significant loci. Moreover, our findings replicate and expand upon previous genetic associations with neuropathies, while at the same time revealing novel signals in genes involved in immunity, neural development, and serotonergic signaling.

The study should be interpreted in the light of the following limitations. First, FinnGen and UK Biobank rely on ICD-coded diagnoses extracted from health records, which may contribute to specificity and accuracy of diagnosis. Misclassification or heterogeneity within diagnostic codes, including the inclusion of varying definitions of neuropathy, could attenuate associations or obscure a subset of the association signals.

Second, while our sensitivity analysis excluded individuals with diabetic neuropathies, residual confounding by comorbid conditions (such as cancer or autoimmune diseases) remains possible, particularly for loci in the MHC region where pleiotropy is common. Disentangling primary genetic associations from secondary effects driven by comorbid conditions needs to be evaluated by future studies.

Third, although the study design included participants of predominantly European ancestry, this limits the generalizability of the associations to other ancestries and may miss associations that cannot be captured by the current cohorts. Future studies in more diverse populations are required to validate these findings and identify additional ancestry-specific risk loci.

Finally, while eQTL and colocalization analyses provided functional associations, they were limited to available tissue datasets, which may not fully capture the relevant biology of peripheral nerves or small fiber neuropathies. Functional follow-up studies, including gene expression profiling in disease-relevant tissues and experimental validation, are needed to clarify the mechanistic roles of implicated genes such as *DIRC3* and *HLA-DQB1*.

In conclusion, we have performed a meta GWAS of neuropathies using FinnGen and UKB, where we found 48 novel loci associated with neuropathies. We found additional evidence for the impact of two of those loci in the genes *DIRC3* and *HLA-DQB1*. Variants in these loci should be considered when studying neuropathies and evaluating risk factors. Furthermore, our data suggest therapies and treatments of comorbid sleep problems and psychiatric traits in handling the health impact of neuropathies. The role of the relatively unexplored gene *DIRC3* should be explored further, particularly in brain tissues, which may yield further insights into the connection between neuropathies and sleep and psychiatric traits.

## Data Availability

All data produced in the present study are available upon reasonable request to the authors

## Acknowledgements

This work was supported by a grant from the Juhani Aho Foundation for Medical Research.

We want to acknowledge the participants and investigators of the FinnGen study. The FinnGen project is funded by two grants from Business Finland (HUS 4685/31/2016 and UH 4386/31/2016) and the following industry partners: AbbVie Inc., AstraZeneca UK Ltd, Biogen MA Inc., Bristol Myers Squibb (and Celgene Corporation & Celgene International II Sàrl), Genentech Inc., Merck Sharp & Dohme LCC, Pfizer Inc., GlaxoSmithKline Intellectual Property Development Ltd., Sanofi US Services Inc., Maze Therapeutics Inc., Janssen Biotech Inc, Novartis AG, and Boehringer Ingelheim International GmbH. Following biobanks are acknowledged for delivering biobank samples to FinnGen: Auria Biobank (www.auria.fi/biopankki), THL Biobank (www.thl.fi/biobank), Helsinki Biobank (www.helsinginbiopankki.fi), Biobank Borealis of Northern Finland (https://www.ppshp.fi/Tutkimus-ja-opetus/Biopankki/Pages/Biobank-Borealis-briefly-in-English.aspx), Finnish Clinical Biobank Tampere (www.tays.fi/en-US/Research_and_development/Finnish_Clinical_Biobank_Tampere), Biobank of Eastern Finland (www.ita-suomenbiopankki.fi/en), Central Finland Biobank (www.ksshp.fi/fi-FI/Potilaalle/Biopankki), Finnish Red Cross Blood Service Biobank (www.veripalvelu.fi/verenluovutus/biopankkitoiminta), Terveystalo Biobank (www.terveystalo.com/fi/Yritystietoa/Terveystalo-Biopankki/Biopankki/) and Arctic Biobank (https://www.oulu.fi/en/university/faculties-and-units/faculty-medicine/northern-finland-birth-cohorts-and-arctic-biobank). All Finnish Biobanks are members of BBMRI.fi infrastructure (https://www.bbmri-eric.eu/national-nodes/finland/). Finnish Biobank Cooperative -FINBB (https://finbb.fi/) is the coordinator of BBMRI-ERIC operations in Finland. The Finnish biobank data can be accessed through the Fingenious® services (https://site.fingenious.fi/en/) managed by FINBB.

