## Supplementary figures and images for "Genome-Wide Meta-Analysis Identifies Genetic Risk Loci for Mono- and Polyneuropathies in 983,477 Individuals"

### Supplemental Figure 1

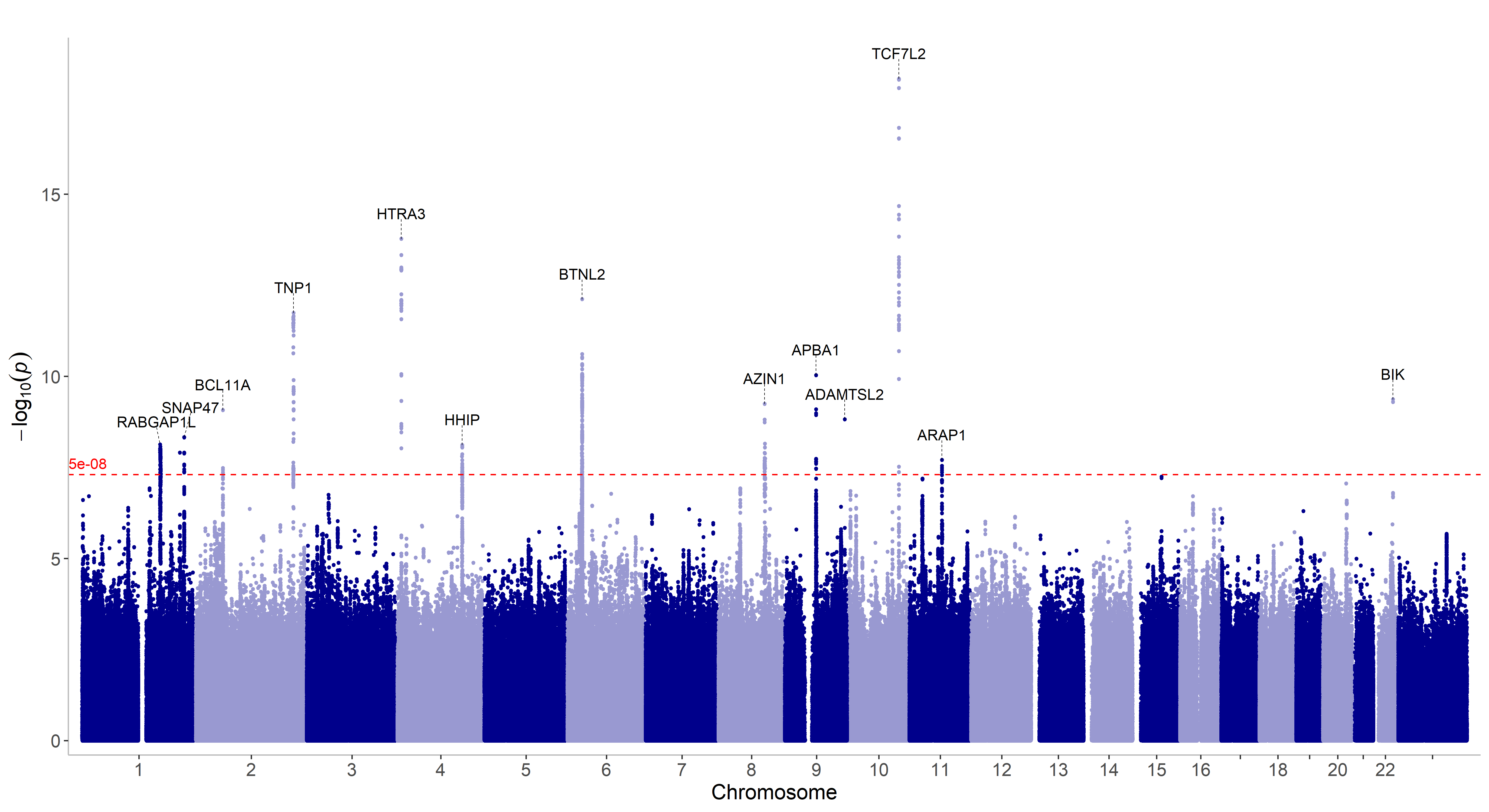
